# A protocol for evaluating digital technology for monitoring sleep and circadian rhythms in older people and people living with dementia in the community

**DOI:** 10.1101/2023.11.10.23298264

**Authors:** Ciro della Monica, Kiran K. G. Ravindran, Giuseppe Atzori, Damion Lambert, Thalia Rodriguez-Garcia, Sara Mahvash-Mohammadi, Ullrich Bartsch, Anne C. Skeldon, Kevin Wells, Adam Hampshire, Ramin Nilforooshan, Hana Hassanin, UK Dementia Research Institute Care Research & Technology Research Group, Victoria L. Revell, Derk-Jan Dijk

## Abstract

Sleep and circadian rhythm disturbance are predictors of poor physical and mental health, including dementia. Long-term monitoring of sleep and circadian rhythms in people living in the community may have great potential for early diagnosis, monitoring of disease progression and assessing the effectiveness of interventions in dementia and other health conditions. Many novel digital technology-based approaches for monitoring sleep and circadian rhythms, aimed at both the consumer and research markets, have recently become available. However, before such technology can be implemented at scale, its performance and acceptability need to be evaluated and compared to established gold standard methodology in relevant populations.

Here we describe a protocol for the evaluation of novel sleep and circadian technology in cognitively intact older adults, some of whom have stable comorbidities, and in people living with dementia (PLWD). In this protocol a range of technologies is tested simultaneously first at-home during a one- to two-week period and subsequently in a clinical research facility in which gold standard methodology for assessing sleep and circadian physiology is implemented.

We emphasize the importance of assessing both nocturnal and diurnal sleep (naps), valid markers of circadian physiology, and that evaluation of sleep and circadian technology is best achieved in protocols in which sleep is mildly disturbed and in populations and settings that are relevant to the targeted use-case. We provide details on the design, implementation, and challenges of this sleep and circadian technology evaluation protocol along with examples of the datasets that have been generated in both cognitively intact older adults and PLWD. We place our protocol in the context of existing literature and highlight the benefits that our methodology has over previous approaches.

**Author Summary:** Sleep and circadian (24-hour) rhythms are frequently disrupted in various physical and mental health conditions including dementia. These disturbances may merely be an outcome of the disease process or be drivers of disease progression. Either way, the ability to monitor sleep and circadian rhythms long-term and at-home could be highly beneficial for monitoring health status, early identification of decline, and implementation of interventions as well as monitoring the effectiveness of interventions. A number of new digital health technologies are now available that could allow non-invasive, non-intrusive, long-term monitoring of sleep and circadian rhythms at home. However, few of these have been evaluated against gold-standard measures in a relevant population. Here we describe our approach to simultaneously evaluating multiple types of sleep and circadian technologies against standard methodology, both at-home and in the laboratory, in a group of older adults with and without dementia. We discuss the feasibility of this protocol, provide examples of the data that were obtained, the data analyses and visualisations that can be applied, and explain the benefits of our approach.

## Introduction

### The need for technology to monitor sleep and circadian rhythms longitudinally

Sleep and the circadian system are important contributors to well-being and both physical and mental health [1–4]. Disruptions to sleep or the circadian clock in ill health may be a predictor of and/or contributor to disease progression as well having a negative impact on quality of life. Much of our knowledge in this area is based on self-report, cross-sectional studies or short-term laboratory studies. The capacity to unobtrusively monitor sleep-wake cycles, circadian rhythms, and other health indicators over long periods of time at home offers opportunities: a) to monitor disease progression and associated clinical outcomes, b) for early detection of decline and implementation of appropriate action, c) to increase understanding of the relationship between sleep/circadian physiology and clinical symptoms/intervention/recovery/response. In addition, longitudinal monitoring of sleep and the circadian system within an individual may facilitate the development of personalised interventions to improve clinical symptoms and quality of life.

The timing, duration and quality of sleep is regulated by an interaction of two well- characterised biological processes: circadian rhythmicity, and a sleep-wake dependent process which is often referred to as sleep homeostasis [5–8]. The sleep homeostat monitors time awake whereas the circadian system drives 24-hour rhythms in nearly all aspects of our physiology and behaviour. The master circadian clock, the suprachiasmatic nuclei (SCN) of the hypothalamus is synchronised to the 24-hour day by environmental light (e.g.,[3, 9]); in addition, environmental light can also modify our levels of alertness, mood, and performance [10].

People living with dementia (PLWD) and their caregivers are examples of populations that may benefit from monitoring of sleep and circadian rhythms over long periods of time. Disturbances of sleep and circadian rhythms are highly prevalent in dementia and include night-time awakening and wandering, long naps during the daytime, and early or late sleep timing [11–14]. Sleep timing disturbances may vary across various dementias such as fronto- temporal dementia (FTD) and Alzheimer’s disease [15]. In addition, sleep disorders are prevalent in dementia, particularly obstructive sleep apnoea and REM sleep behaviour disorder which are risk factors for dementia and neurodegeneration and contribute to cognitive decline [16–18]. These disruptions not only have consequences for health and social care costs but also affect the quality of life of both the PLWD and their care giver (e.g., [19–22]) and are a major contributing factor to the PLWD being moved into care homes (e.g., [23–25]). These sleep and circadian disturbances may be a consequence of the disease process and, as such, be an indicator of disease progression (e.g., [26]). Alternatively, or additionally, sleep disturbances may actually drive disease progression and thus be a target for intervention to improve the quality of life of both PLWD and their carers. Furthermore, night-to-night variation in aspects of sleep and sleep continuity in particular have been shown to predict day-to-day variation in symptoms in dementia [27]. Longitudinal monitoring of sleep and circadian physiology will benefit our understanding of their interaction with health outcomes in dementia.

### Gold standard assessments of sleep and circadian rhythms: advantages and disadvantages

We can measure sleep in many different ways from simple self-report (e.g., sleep diaries), to increasing complexity at the behavioural (e.g. bed occupancy), and physiological (e.g., EEG, cardiovascular) level. In addition, the classification of sleep can be made at the simple sleep vs wake distinction to the more detailed macrostructure of the different stages of non-rapid eye movement sleep (NREM) stages N1 to N3 and REM sleep, and finally to the microstructure of the electroencephalogram (EEG) signal, as reflected in power spectral density or other EEG measures. From these different levels of measurements we can derive a range of parameters to describe sleep from self-reported sleep quality to the timing of sleep within the 24-h day, total sleep time (TST), sleep onset latency (SOL), wake after sleep onset (WASO), sleep efficiency (SE), spectral power of different EEG frequency bands, and individual EEG events such as slow waves and sleep spindles as well as their phase relationships (e.g., [2]).

The gold standard method of assessing sleep is laboratory-based polysomnography (PSG) which is performed in accordance with guidelines of the American Academy of Sleep Medicine [28]. PSG is a comprehensive overnight physiological assessment including EEG, electrooculogram (EOG), electromyogram (EMG), electrocardiogram (ECG), oxygen saturation (SpO2), respiration effort and airflow, limb movement (EMG), body position, and video recording. The recordings can then be scored to provide a detailed picture of sleep structure and physiology, including the presence of any clinical sleep disorders such as sleep apnoea where, for example, the apnoea-hypopnea index (AHI) from a single night can be used to determine disease presence and severity. In addition, periodic limb movements can be measured to assess for sleep-related limb movement disorder.

To understand the contribution of the circadian system to health and disease it is necessary to be able to characterise its properties which includes: the phase (timing), amplitude (strength), and period (time taken for one complete cycle) of the rhythms.

Traditionally, assessment of phase and amplitude has been achieved by acquiring time series data of gold standard measures (e.g., melatonin, cortisol, core body temperature) in highly controlled laboratory conditions (i.e., dim light, continual wakefulness, controlled posture, controlled calorie intake) [29]. More recent approaches utilise machine learning or mathematical models to extract features from only a few samples of high dimensional data e.g., transcriptomics, metabolomics, or longitudinal simultaneous recordings of light exposure, activity, and physiology [29]. However, many of these approaches have yet to be tested and validated in different populations or under different sleep/wake, light/dark schedules.

These gold standard approaches can provide a detailed assessment of an individual’s sleep and the phase/amplitude/period of their circadian system. However, these assessments: a) have high associated cost due to the requirement for participants to be supervised in a laboratory environment by appropriately skilled staff, b) impose a high level of burden on participants due to amount of equipment that needs to be worn and needing to travel away from home to a laboratory setting, c) can be invasive if blood samples are collected, d) require technical analysis skills e.g., for scoring the PSG recording, and e) unrepresentative of normal individual sleep patterns due to first night effects and novel controlled surroundings. Moreover, a single PSG recording and single melatonin profile in the laboratory only provides a snapshot of an individual’s sleep/circadian physiology and behaviour. Whilst these gold-standard approaches are essential for diagnoses or research studies they do not provide measures of ecological validity within an individual’s home over an extended period of time.

### Technology for monitoring sleep and circadian rhythms at home: current approaches

New digital health technology to monitor sleep/circadian behaviour and physiology at home is constantly emerging on the consumer and research markets. Consumer monitoring devices are designed to appeal to the general public in terms of cost, appearance and the type or level of information that they provide. These devices can potentially provide behavioural level data, including bed occupancy and activity, as well as more detailed physiological signals from which sleep stage, heart rate and breathing rate can be provided. However, as these are consumer rather than medical devices, no particular level of quantitative performance is mandated or guaranteed. Nonetheless, the ability to cost-effectively monitor sleep and circadian rhythms in an individual’s own home offers several advantages. Firstly, the assessments are made in a natural environment which allow the influence of daily activities/behaviours and local environmental factors, including light and temperature, on sleep and circadian rhythms to be assessed. In fact, many devices also measure environmental variables including light, noise, and air quality. Secondly, assessments can be made longitudinally so it is possible to determine how sleep/circadian rhythms vary over time and investigate how these longitudinal variations associate with changing behaviours or symptoms. Finally, the capacity to monitor sleep within a home environment may be beneficial for those suffering from sleep disorders (e.g., sleep apnoea) where the technology may assist with diagnosis, clinical monitoring, and/or the delivery of, and monitoring the efficacy of interventions.

Longitudinal assessments of circadian rhythms in the field, to date, have taken three approaches: 1) measuring rest activity patterns with wrist-worn actigraphy in conjunction with sleep diary, and using sleep timing as a proxy for the timing of the clock, 2) assessment of circadian phase through sample collection and measurement of melatonin or its metabolites at defined intervals, 3) combining light and activity measurements with mathematical models to predict circadian phase [29].

Actigraphy records limb movement activity (accelerometery) and then uses a proprietary algorithm to process this movement data to estimate whether an individual is awake or asleep for each 60 second epoch, and subsequently derive sleep measures including TST, SOL and WASO. In addition, non-parametric analysis can be conducted to determine a range of related variables: inter-daily stability (IS), a measure of day-to-day consistency of activity patterns, intra-daily variability (IV), a measure of how much activity varies within a 24-hour period, 10 h of highest activity (M10), 5 h of lowest activity (L5), and relative amplitude (M10: L5). However, the use of the timing of sleep/rest periods as an estimate for circadian phase is not advisable [3]. The relationship between the circadian clock, as indexed by melatonin, and sleep timing varies in both healthy individuals [30] but also in different mental health conditions [3]. Consequently, sleep/rest timing is not a reliable indicator of the timing of the internal clock. The phase relationship between sleep and circadian rhythms are relevant, such that it for example predicts whether or not late sleep timing (eveningness) associates with depressive symptoms [4].

Field assessments of the gold standard marker of circadian phase, i.e., melatonin profiles, are challenged by the fact that melatonin is sensitive to exogenous factors including environmental light and posture such that participants need to follow a controlled protocol. Collection of saliva samples, under dim light whilst seated, at 30 min intervals in the 3 – 4 hours before habitual bedtime allows the dim light melatonin onset (DLMO) to be determined as a marker of circadian phase. Implementation of technology, including containers that track when salivettes are removed for sampling, have allowed the development of home protocols that have been validated, in a within-subject design, against DLMO collected in the laboratory [31]. An alternative approach that is less restrictive for participants is 48-hour urinary collections to measure the urinary metabolite of melatonin, 6-sulphatoxy-melatonin (aMT6s). This methodology has been used successfully in both blind individuals and those living with schizophrenia, who frequently suffer circadian and sleep disruption, to track circadian phase over several weeks (e.g., [32–34]). It should be noted that this approach may cause burden to participants and, as the circadian parameters are computed from a rhythm derived from samples collected over 4 – 8-hour bins, the markers may not have sufficient resolution to detect small but relevant changes in circadian phase. More recently, statistical and mathematical models for the interaction between the circadian system, sleep homeostat and environmental light exposure approaches have been successfully applied to wearable data to predict circadian phase [35–37].

### Evaluating technology: the issues

The main issues with novel technology are: 1) lack of evaluation against gold standard measures such that the device output cannot be verified, 2) consumer device hardware and algorithms are constantly being updated which means that any evaluation that has been performed may be rapidly out of date, 3) if evaluation studies are performed then they are typically in young, healthy individuals for a habitual time-in-bed period where sleep efficiency is high and where rest/activity rhythms are robust and regular.

These issues of evaluation studies means that the device performance may not translate to situations of disturbed sleep/circadian rhythms or in clinical/older populations who may benefit from long-term use of the devices. For example, sleep undergoes well-characterised changes with age, but also in dementia, at both the macro- and microstructure level. These macro changes include changes in sleep timing, shortened or extended nocturnal sleep duration/time in bed, increased number of nocturnal awakenings and time spent awake during the night, decreased slow wave and REM sleep, and increased frequency of daytime naps (reviewed in [38]). At the microarchitecture level, with age and in dementia there are reductions in slow wave activity (SWA) in NREM sleep, particularly in the prefrontal cortex and during the first NREM cycle of the sleep episode, as well as a decreasing number of sleep spindles and reductions in REM sleep (reviewed in [39–41]).

This alteration in both the sleep signal and sleep continuity with age and in dementia implies that sleep monitoring devices that perform well in young people may not do so well in older people or in people living with dementia. Nevertheless, there is a lack of evaluation studies in particular in PLWD [42]. For example, a recent systematic review of the validity of non-invasive sleep-measuring devices aimed at assessing their future utility in dementia, was not able to identify any studies in people with mild cognitive impairment or Alzheimer’s Disease [43].

In addition to changes in sleep, aging is associated with changes in the circadian system in terms of its timing, amplitude, and relationship with sleep (e.g., [44]) as well as changes in light exposure, crucial for stability and robustness of the clock, due to changes in photic sensitivity (e.g., [45]) and the lived light environment (e.g., [46]). As such, circadian technologies may not perform as well in older individuals.

The ‘International Biomarkers Workshop on Wearables in Sleep and Circadian Science’ held at the 2018 SLEEP Meeting of the Associated Professional Sleep Societies identified that the main limitation of large-scale use of novel sleep and circadian wearables is the lack of validation against gold-standard measures [47]. The workshop formulated guidelines for validation and confirmed that: PSG is the only valid reference for TST and sleep staging; PSG sleep records should be scored using current AASM guidelines; PSG sleep records should be double scored to minimise bias. Devices differ in the level at which they classify sleep-wake, from binary (sleep or wake) to four stages (wake (W), REM, light sleep (LS), deep sleep (DS)) to full AASM (W, stage 1 NREM (N1), stage 2 NREM (N2), stage 3 NREM (N3), REM) scoring. The ability of a device to over or underestimate sleep and wake will depend on its sensitivity (ability to correctly classify sleep epochs), specificity (ability to correctly classify wake epochs), and accuracy (proportion of all epochs correctly detected) (reviewed in [47, 48]). These factors will depend on the physiological variables and classification system used to determine sleep and wake. For example, according to traditional performance measures, actigraphy tends to have high sensitivity and accuracy but low specificity [49]. Despite this, actigraphy is considered a valuable tool for long-term monitoring of rest/activity patterns in clinical populations [50].

Although it is crucial to evaluate the performance of a device against gold-standard measures, perhaps even more important is to assess its performance longitudinally in the real world where it will be used in both older adults and PLWD. Simultaneously, to increase our understanding of sleep and circadian rhythms in the real-world and their interaction with disease processes, it is important to monitor relevant environmental variables that may impact sleep or the circadian system, in particular environmental light which is critical for synchronising the circadian system (e.g., [9]) as well as modifying our levels of alertness, mood, and performance. In addition to assessing performance, it is also critical to determine that the device is cost-effective and scalable, and to assess the acceptability of novel sleep/circadian monitoring devices in the end-user group. This will include assessing comfort, burden, obtrusiveness, and how easy the device is to use.

Here we describe our approach to evaluating novel wearable and contactless sleep, circadian and environmental monitoring technology in community-dwelling adults both at home and in the laboratory using multiple devices simultaneously against accepted standard measures. We use this approach in cognitively intact older adults as well as PLWD and their caregivers. Here, we provide some examples of the results from cognitively intact adults and PLWD. Whilst our protocol focuses on technology that can be used in PLWD for long-term monitoring at home, this approach could be applied to many technologies and populations.

## Materials & Methods

Our approach was to first conduct a study in cognitively intact older adults to assess the performance and acceptability of a range of devices before utilising our initial findings to design a feasibility protocol to implement selected technology in PLWD and their caregivers. Both studies used the same approach of multiple days recording at-home followed by an in-laboratory, residential overnight session. The study in cognitively intact adults is completed, and some of the results have been published [51, 52] and the study in PLWD and their caregivers is ongoing.

### Study Design

#### Ethical approval

The study in cognitively intact older adults received a favourable opinion from the University of Surrey Ethics Committee, and the study in PLWD and their caregivers from an NHS ethics committee (22/LO/0694). The protocols are conducted in accordance with the Declaration of Helsinki and guided by the principles of Good Clinical Practice. Written informed consent is obtained from participants before any procedures are performed. All participants are compensated for their time and inconvenience. Within the participant information sheet, it is clearly stated that all personal data is handled in accordance with the general data protection regulations (GDPR) and the UK Data Protection Act 2018. In addition, it is explained that anonymised non-personal data may be transferred to the manufacturers of the devices being tested if they need to process the data. The manufacturer can only access anonymised data that details the serial number of the device, the date of recording, and the signals recorded on that specific date. The manufacturers may use this anonymised data in their continual assessments of the performance of their device.

#### Participants

For an evaluation study, the study population selected must ensure that the findings are relevant to the use case. For our initial protocol in cognitively intact older adults, the eligibility criteria for participation in the protocol were designed to maximize the plausibility of relevance for the PLWD population. As such, the target population consisted of cognitively intact, independently living, non-smoking, men and women aged between 65 – 85 years. Participants had to consume ≤ 28 units of alcohol per week and were current non- smokers. Those with stable controlled medical conditions were included to ensure a representative heterogeneous population for this age range. This approach is in contrast to the stringent criteria, in relation to health conditions and medication, applied for most clinical trials.

For our protocol in PLWD and their caregivers, the inclusion/exclusion criteria for PLWD include: age range of 50 – 85 years, a confirmed diagnosis of prodromal or mild Alzheimer’s disease, an S-MMSE (Standardised Mini Mental State Examination, [53]) score ≥ 23, living in the community and, if taking medication for dementia, being on a stable dose for three months prior to recruitment. Individuals who have an unstable mental state, severe sensory impairment, active suicidal ideation, or being treated for terminal illness are not included. PLWD can participate in the study by themselves, or their carer/family support/friend may also enrol as a ‘study partner’ participant. These study partners must be ≥ 18 years, have an S-MMSE score ≥ 27, and must have known the PLWD for at least six months and be able to support them in their participation. Study partners complete the same procedures as the PLWD.

Cognitively intact older adults were recruited via our Clinical Research Facility database where potential participants have registered and consented to be contacted about ongoing research. PLWD and their study partners are recruited in collaboration with local NHS trusts via memory services. All participants undergo an initial telephone interview and subsequent in-person screening visit to determine their eligibility to take part in the study.

At the screening visit for cognitively intact older adults, following informed consent, participants completed a range of assessments including measurement of height, weight and vital signs (body temperature, heart rate, respiration rate and blood pressure), self-reported medical history, and completion of baseline questionnaires: Epworth Sleepiness Scale (ESS) [54] (> 10 indicates excessive daytime sleepiness), Pittsburgh Sleep Quality Index (PSQI) [55] (> 5 indicates a sleep disorder), Activities of Daily Living Questionnaire (ADL) [56] and International Consultation on Incontinence Questionnaire – Urinary Incontinence (ICIQ-UI) [57].

At the screening visit for PLWD (and study partners, when applicable), and following informed consent, the participants complete standard assessment tools which are frequently used in this population (i.e. sMMSE [53]) Hospital Anxiety and Depression Scale (HADS) [58], quality of life in Alzheimer’s Disease (QoL-AD) [59] as well as the PSQI [55], and medical history questionnaire. In addition, vital signs are recorded as well as height, weight, and BMI. PLWD and their study partners also complete additional questionnaires either at this visit or during their overnight sessions: ESS, ADL, ICIQ, National adult reading test (NART) [60], Berlin questionnaire to assess for sleep apnoea [61], Horne-Ostberg questionnaire [62] to assess time of day preference, and their education level is documented. For all participants, their general practitioner is informed of their participation.

#### Technology evaluated

Within our protocol we have categorised the technology according to how it is used or what it is monitoring: a) wearable devices that are placed on the body (e.g., wrist, head), b) nearable/contactless devices that are placed near the individual (e.g., bedside or under the mattress) to detect physiological or behavioural signals, c) environmental monitoring devices (e.g., light, temperature), d) use-able devices that the participant interacts with (e.g., electronic tablet for cognitive testing), e) video monitoring that provides information about the individual and the environment.

The technology assessed in the current study was selected on the basis of: a) previous evaluation studies, b) inclusion in comparable studies, c) regulatory status, d) cost, e) potential acceptability to PLWD. We included both research grade and consumer directed devices. The number of devices tested simultaneously took account of potential burden on participants and ensured that adequate signals could still be obtained when multiple devices were worn (appropriately positioned). Cognitively intact participants wore no more than four wrist devices (two per arm) at any one time whereas in PLWD no more than two were worn. The technology used in each protocol is shown in Table 1.

**Table 1.**
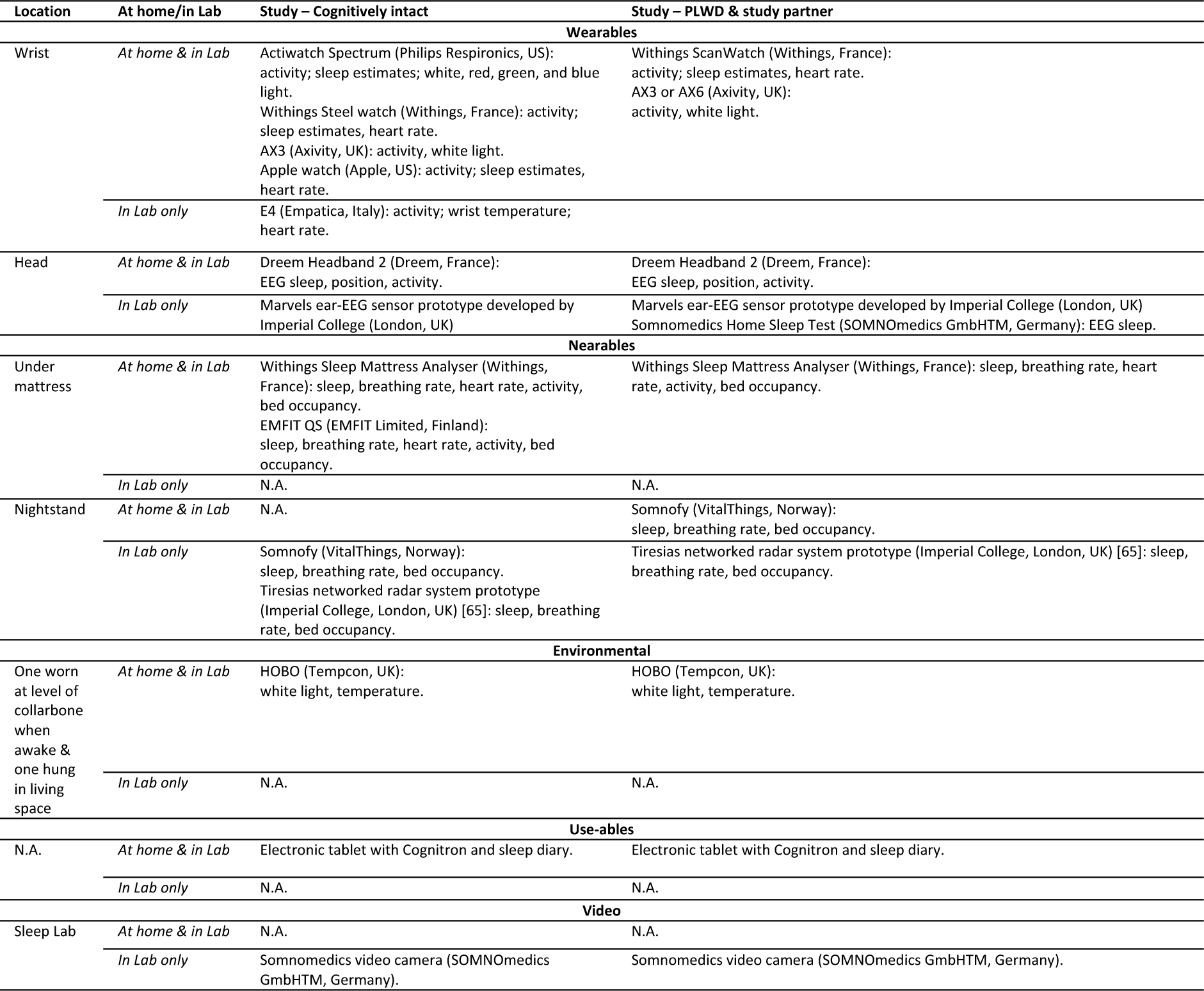
Commercially available and research-grade devices used in the study protocols.

#### Study Protocol

These studies are designed to assess the performance and acceptability of a range of devices simultaneously, firstly longitudinally at-home (7 – 14 days) with actiwatch/sleep diaries as a reference point and then in an overnight laboratory session with concurrent gold standard video-PSG. The AASM recommends that actigraphy is always accompanied by completion of sleep diaries to allow optimal interpretation of the actigraphic data [63] and that data is collected for a minimum of 72 hours to 14 days [50]. The concept of the study is shown in Figure 1 and a schematic diagram of the protocol is shown in Figure 2.

**Figure 1.**
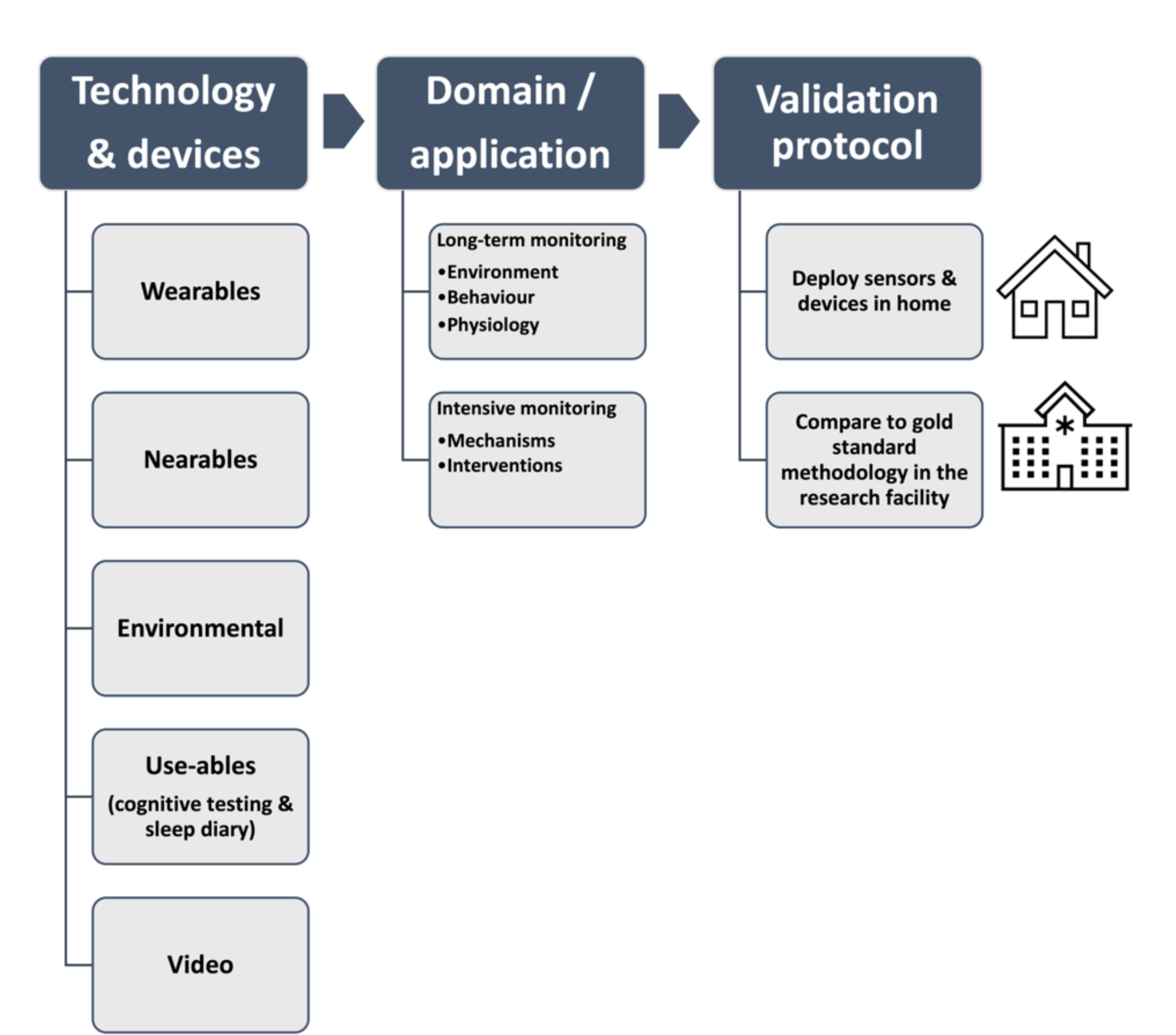
Overview of the concept of the protocol and categories of the devices included, the data and application domain, and the study design.

**Figure 2.**
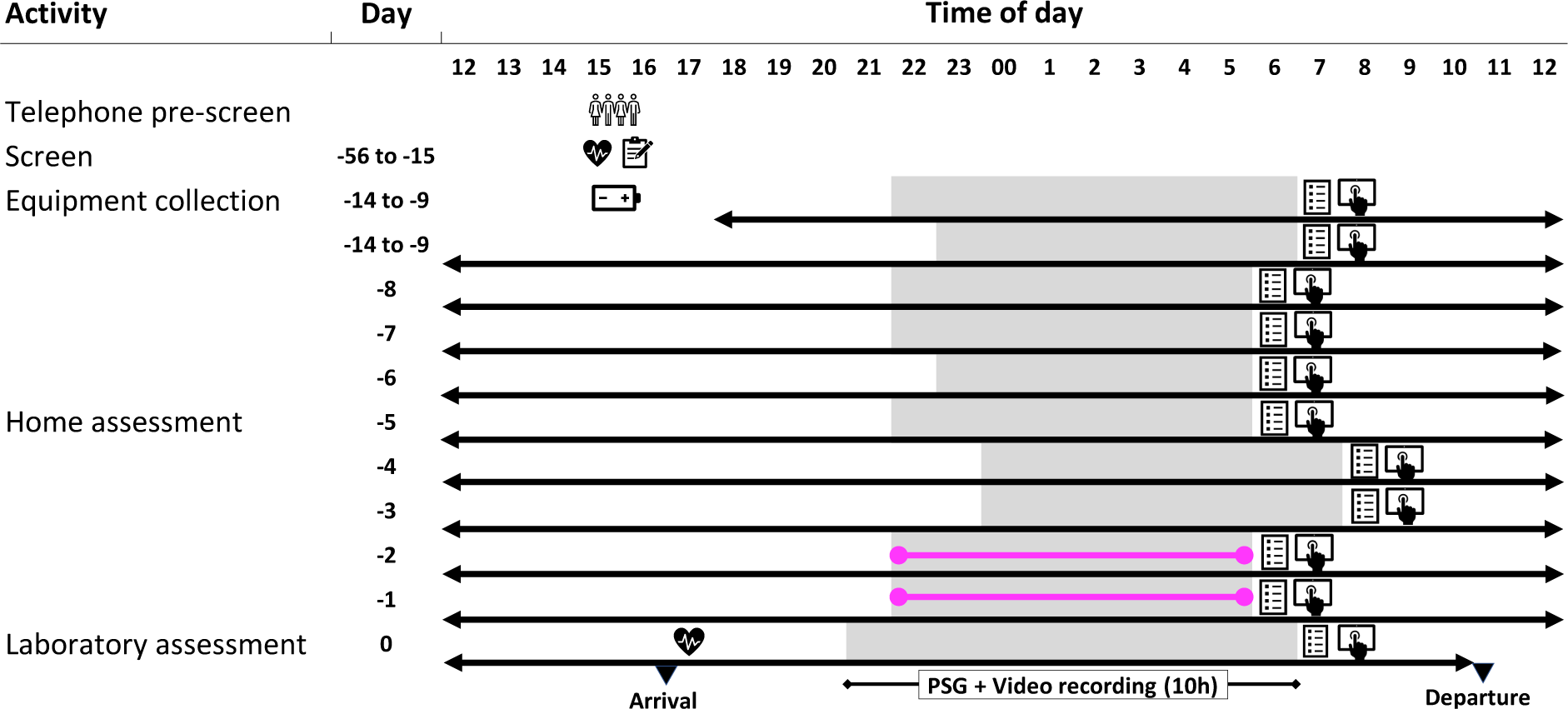
Schematic diagram of the protocol design. The people symbol indicates when the telephone pre-screening assessment was conducted. The clipboard symbol indicates completion of questionnaires, the heart symbol when vital signs were measured, and the battery symbol indicates when participants were trained to use the technology. Grey bars indicate sleep periods and white bars indicate wake periods. The symbol of a hand using an electronic tablet indicates completion of the cognitive test battery, and the questionnaire is completion of the sleep diary. The pink horizontal lines indicate when an EEG device was used at home to measure sleep physiology. The black horizontal line indicates the use of wearables and nearables throughout the at-home period.

##### Longitudinal monitoring at-home

Participants are provided with a range of technology to use in their home to monitor their sleep/wake patterns and environmental light exposure (see Table 1 for list of devices and variables measured). The technology is either installed by the participants themselves or researchers may go to the participants’ homes to assist them. The wrist-worn/collarbone-worn devices are worn continually and participants are requested to complete a log whenever they remove them to record times and reason for removal. The EEG wearables are only used for one or two nights and the nearables are left *in situ* throughout. Participants are requested to complete a modified version of the Consensus Sleep Diary-M [64] (electronically or on paper) on a daily basis to record subjective information about their sleep patterns, sleep quality, daytime napping as well as alcohol and caffeine consumption. In addition to the standard questions, participants are asked to provide further details about their daytime naps (what time, duration, where and why they napped) and nocturnal awakenings (for each awakening, what time they awoke, how long it took to fall asleep, if they left the bed, and if so, what time). Participants are requested to complete cognitive assessments one to two hours after waking each day on an electronic tablet.

##### Overnight laboratory session

The session is ∼24 hours in duration and participants are required to arrive in the afternoon and remain at the Research Centre, which hosts the UKDRI clinical research facility at Surrey, until the following day. Upon arrival, participants’ vital signs are measured, and continued eligibility assessed. The devices that have been used at home are downloaded, reset, and provided back to the participants with any additional devices only used in the laboratory. During their stay, participants also have assessments of their gait and postural stability made using video and radar technology.

During the laboratory session, participants have an indwelling cannula sited for collection of regular blood samples at 3-hourly intervals (including overnight) for 24 hours to assess time of day variation in biomarkers. The samples are processed and are analysed for levels of melatonin, as a gold-standard marker of the circadian clock, as well as biomarkers of dementia e.g., neurofilament light (NfL), phosphorylated tau (p-tau), amyloid-beta (Aβ40, Aβ42 (e.g., [66–68]). In addition, participants collect urine for 24 hours in four-hourly intervals (eight hours overnight) for measurement of aMT6s.

Following dinner, participants are equipped with all the electrodes and sensors required for a clinical video-polysomnographic (PSG) recording using AASM compliant equipment and montage. The PSG equipment is the Somnomedics SomnoHD system with Domino software (v 3.0.0.6, sampled at 256 Hz (SOMNOmedics GmbHTM, Germany) and we use an American Academy of Sleep Medicine (AASM) standard adult montage. Participants can also have a wearable EEG device fitted for concurrent EEG recording and contactless sensors are positioned for overnight recordings. Prior to the start of the PSG recording, participants are asked to lie on the bed in different supine poses (prone, supine, right, left, seated) and recordings are made with video and radar technology to assess the ability of the radar technology to assess physiology in different poses.

The protocol takes advantage of the ‘first night effect’ and an extended period in bed to create a model for mildly disturbed sleep [69]. Participants are required to be in bed for a 10 h recording period that is determined on the basis of their habitual time in bed period (HTiBP). For example, for HTiBP < 8 h, the recording period starts one hour earlier than habitual bedtime; for HTiBP > 10 h, the recording starts at habitual bedtime. For those with 8 h ≤ HTiBP ≤ 10 h the recording start time is determined as: Habitual Bedtime – [0.5*(10- HTiBP)]. This extended period in bed is used to ensure that the recordings include periods of quiet, recumbent wake to determine if the devices can distinguish quiet wake from sleep. Participants select their own lights off/on times and around these times are permitted to conduct quiet, sedentary activities e.g., watching movies, reading. Overnight recordings are performed in individual, environmentally controlled bedrooms or in our bespoke bedroom facility that has double occupancy or adjacent room access for PLWD and their study partners.

Upon awakening, participants are requested to complete their sleep diary and cognitive test battery as well as a questionnaire about device acceptability. The questionnaire asks participants to rate comfort (1 – very uncomfortable to 7 – very comfortable), ease of use (1 – very difficult to 7 – very ease), and to record any problems. Prior to discharge, vital signs are taken and for cognitively intact older adults the S-MMSE was administered.

#### Device and data management

The flow of data acquisition and data management is shown in Figure 3.

**Figure 3.**
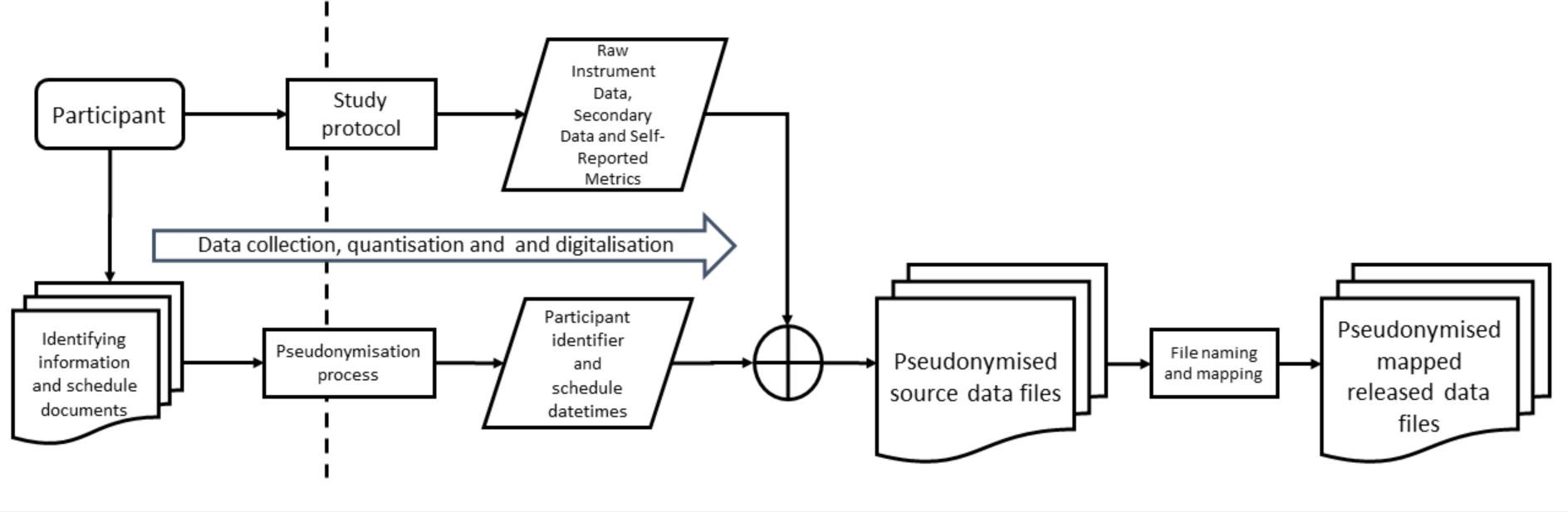
Overview of data acquisition and data management.

##### Device allocation

Eligible participants are enrolled into the study and have a set of devices allocated to them. All devices and systems have unique identifiers, with a one-to-one allocation of: a) device code to participant for wearable/contactless devices used at home or in the lab and b) device to location to participant for devices installed in the laboratory (e.g., floor sensor data is recorded for a specific lab room, and the room is allocated to the participant). All devices are mapped to an operation schedule which means that data are collected during each 24-hour period from 12 noon to 12 noon the following day unless the device has continuous recording.

##### Device set-up and synchronisation

To be able to directly compare the performance of different devices it is essential that they are time synchronised. All network device clocks are synchronised to a Network Time Protocol (NTP) server. Commercial standalone systems are synchronised through the respective software applications used to set up the device recordings.

##### Data acquisition

The devices used are either battery powered logging devices or are directly connected to power and either store data locally or are wi-fi enabled and transmit data to the secure cloud servers of the manufacturers. Participants are provided with an independent Wi-Fi 4G gateway for device connection.

Upon arrival at the laboratory, all devices are collected from participants for: a) download of data and confirmation of power levels for battery powered devices, b) reconfiguration of connected devices to connect to local Wi-Fi connections. Devices to be used during the laboratory session are returned to the participants.

At the end of the lab session all logging devices are connected to the relevant secure system and source data files extracted and moved to a location based on the participant, day(s) and device for that data. For the online server-based systems, these are synchronised, and data is then extracted and placed into the relevant source data file system.

##### Data mapping and file name convention

All data recorded as a source file, or in a source system is mapped to a named file and location based on study specific parameters e.g., Study Name (required), Device code (required), Test/Data/Measure (Optional), Participant or group (required) Visit (required) Study day/night (required). Access to this Research Data Store (RDS) is strictly controlled in accordance with Information Governance procedures for the specific use of the study protocol owners for analysis.

### Data processing and analysis

To evaluate the accuracy and reliability of the focus technology (devices being evaluated) in measuring sleep, we compare it to a standard reference technology. For the at-home recordings, the comparative standard measure against which all other technologies are evaluated is the combination of the Actiwatch-spectrum (AWS) and Consensus Sleep Diary (cognitively intact adults) or AX3 and Consensus Sleep Diary (PLWD and study partners). For the in-laboratory session, the PSG is considered the gold-standard measure [28, 50, 70] for all participants. The PSG recordings are scored in 30 second epochs (in accordance with AASM guidelines) by two independent scorers and a consensus hypnogram is generated.

AHI is determined using the AASM criteria for scoring apnea/hypopneas where there is > 3% drop in oxygen saturation and/or an arousal.

The focus technology is evaluated against the standard reference technology (AWS + sleep diary or PSG) for its ability to estimate sleep summary measures (e.g., TST, SOL). In addition, in the laboratory, the focus technology is assessed for its epoch-by-epoch (EBE) concordance with the PSG hypnogram. Sleep summary measures are obtained from processing data using local individual proprietary software for battery logging devices or via raw signals being uploaded to a cloud-based server for scoring by a proprietary machine-learning based algorithm. Our approach to data analysis is depicted in Figure 4.

**Figure 4.**
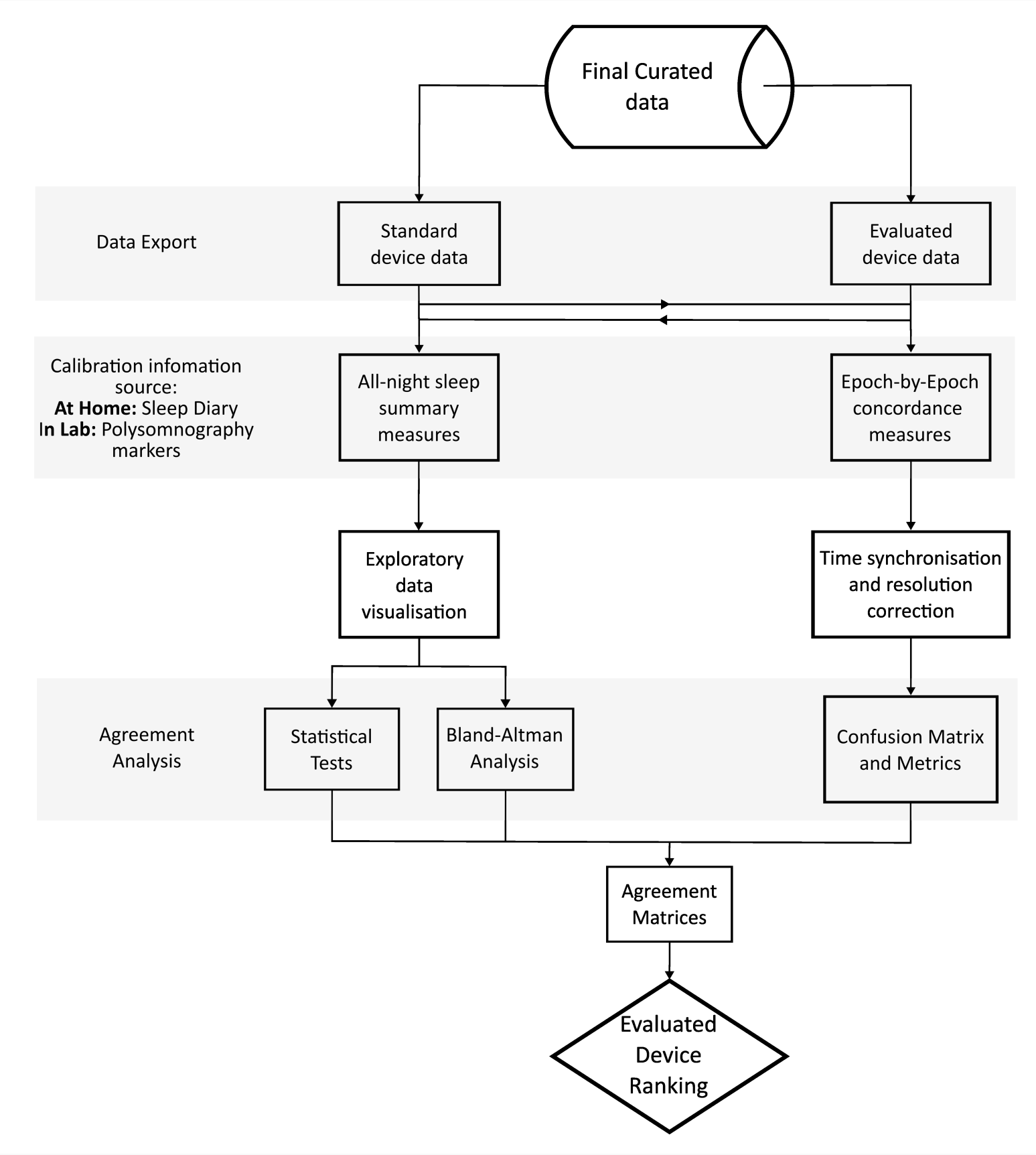
Overview of process for evaluating consumer-grade technology against standard device.

### Sleep summary measures

The sleep summary measures can be grouped into two categories: 1) sleep/wake measures e.g., TST, SOL, WASO, and SE, and 2) sleep stage duration measures. The sleep stage duration measures vary depending on the level at which the focus device classifies sleep-wake i.e., binary, four stages or full AASM. The interval over which the sleep summary measures are calculated (analysis period) is either automatically set by the device algorithm or can be manually set using either the sleep diary reported times of attempted sleep and final awakening (standard for at-home recordings) or the lights off period or total recording period (standard for in-laboratory recording) [28, 50]. The analysis period chosen can have a substantial impact on the summary measures calculated and the performance of the device when compared to home/laboratory standards [51].

All of the focus technologies generate summary measures automatically using the device algorithm determined analysis period. The primary analysis is performed using these automatic summary measure estimates. However, for completeness, the summary measures can also be manually calculated.

A number of data visualisations (scatter plots, box plots, QQ plots, etc.) are performed to check the distribution of the data and for the presence of outliers. Further statistical tests are performed to check the normality of the data. For the agreement estimation, Bland-Altman analysis is performed and bias, limits of agreement and minimum detectable change are estimated. Other metrics that can be computed include for example Pearson’s correlation, consistency intraclass class correlation (ICC), effect size (Cohen’s D) and mean absolute percentage error (MAPE) [71, 72]. To rank the devices, the agreement matrices containing the sleep measure accuracy metrics are created.

### Epoch-by-epoch (EBE) concordance

At-home recordings: the resolution of the focus technology’s hypnogram is reduced to binary sleep/wake classification to match the AWS which is used as the comparative standard measure. The analysis window is set between 18:00 h to 12:00 h and all common periods of sleep/wake timeseries are evaluated.

In-laboratory recordings: the PSG hypnogram resolution is reduced to match the levels of sleep stage output by the device (e.g., N1+N2 = light sleep (LS) and N3 = deep sleep (DS)) to allow direct comparison. Only valid pairs of epochs between PSG and device are used for the concordance analysis. The analysis window is set as the total recording period (∼10 hours).

The EBE concordance metrics for the devices are estimated from the confusion matrices constructed. The concordance metrics used for the analysis of all the different sleep stage levels include, sensitivity, specificity, accuracy, Matthew’s correlation coefficient (MCC) and F1 score. Similar to the sleep summary measures analysis, an agreement matrix for EBE concordance is created using MCC. MCC is preferred to other concordance metrics since it accounts for class imbalance commonly encountered in hypnogram data and is a better alternative to the metrics such as kappa or its variants [73]. The final device ranking is created using the summary sleep measures and EBE agreement matrix. Furthermore, the effect of participant characteristics such as age, sex, BMI, AHI and other confounding factors on the device accuracy and reliability are also explored.

### Environmental measures

Characterising the environment, in particular light, is crucial to understanding sleep/circadian physiology in the real-world and in different disease states. In the current studies, light exposure patterns were assessed by both static and worn devices which recorded lux values at one-minute intervals. One wearable (which measures white, red, green and blue light) was worn on the wrist and one was clipped on clothing near the collarbone; the static device was placed in the room of the home where the participant spent the majority of their time. During data visualisation, imputation was performed for any periods during which the participants were awake, but the measured light levels were 0 lux, which could be due to the sensors being accidentally covered. The imputation consisted of replacing these zero values with the median value from the preceding and succeeding 30 min. This was calculated across all available days of data for each participant separately.

The consistency of measurements between devices was assessed by performing correlations between the lux values obtained by the wrist worn and collarbone-worn devices.

### Quality assurance and mitigating issues Troubleshooting

To maximise data completeness and quality, during the study conduct, data acquisition from wi-fi enabled devices is monitored daily and participants can be contacted if any issues arise. Participants are also able to contact the researchers 24/7 with any issues or concerns. Some potential issues that may arise and their mitigations are presented in Table 2.

**Table 2.**
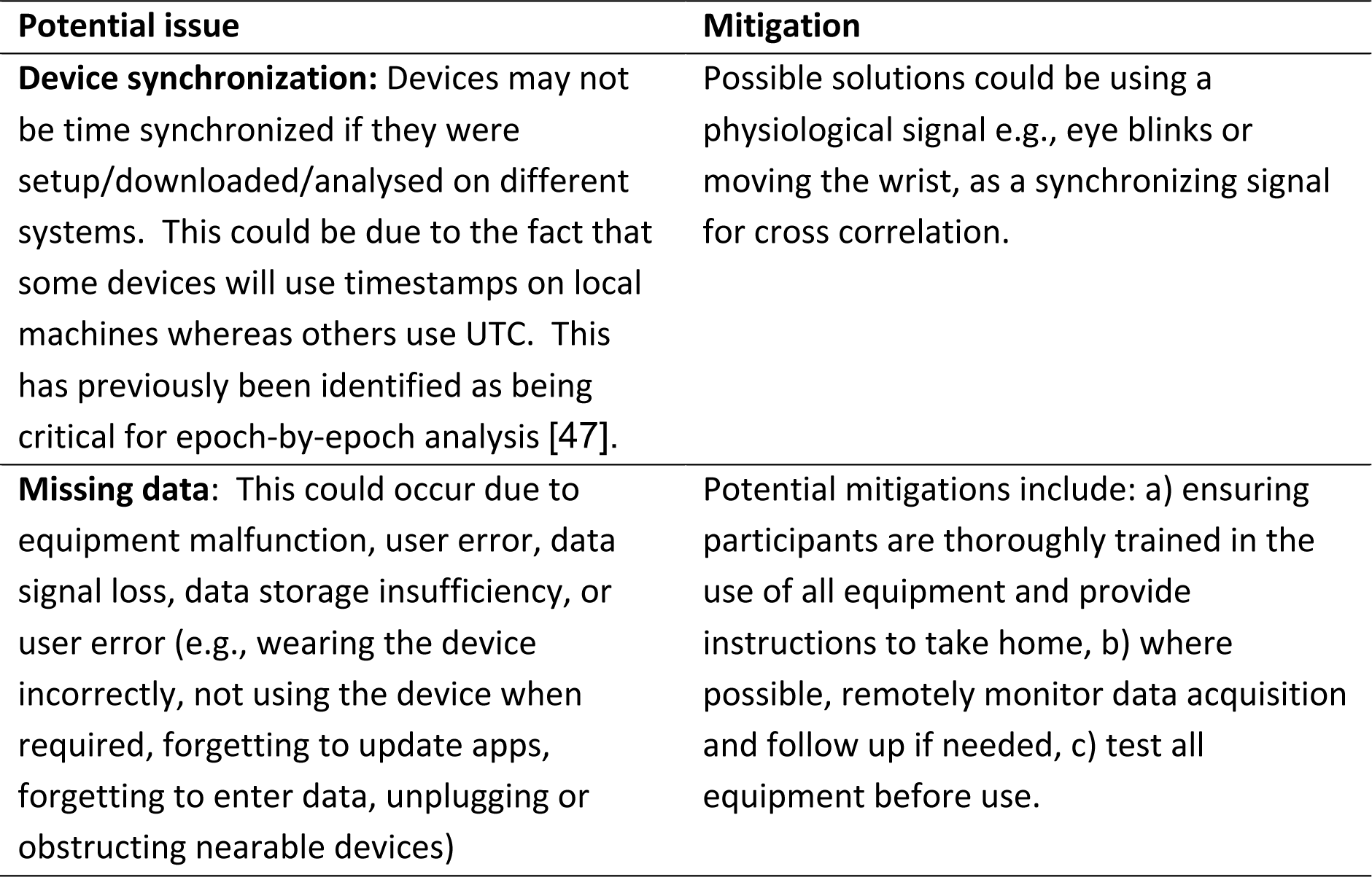
Device evaluation studies: potential issues and mitigations.

### Expertise needed to implement the protocol

These studies require a team of trained personnel to ensure participant safety and well-being as well as data quality and integrity including troubleshooting issues with devices. This level of support is required from the point of consent, throughout the at-home data collection and for the overnight laboratory session. In addition, specialised and competent staff are required for collecting blood samples, PSG instrumentation, PSG recordings, PSG scoring, and data analysis.

## Results

### Participant characterisation

For our initial protocol, 45 out of 46 cognitively intact participants were screened and deemed eligible for the study and, of these, 36 participants were enrolled in the study and 35 participants (14 female: 21 male) completed the study. They had a mean age of 70.8 ± 4.9 years (range: 65 – 83 years) and a mean BMI of 26.7 ± 4.7 kg/m^2^ (range: 20 – 40) which is representative of the UK population in this age bracket [74]. Current co-morbid stable medical conditions were reported by 40% of participants, including type 2 diabetes, arthritis, and hypertension, and 26% of participants were on prescribed medications. The participants had the following scores (mean ± SD) from the baseline questionnaires: a) PSQI: 4.1 ± 2.1, b) ESS: 3.6 ± 2.5, c) ADL: 7.9 ± 0.2, d) ICIQ: 1.0 ± 1.7, e) S-MMSE: 28.7 ± 1.4. From the laboratory session, it was identified that 89% of participants had apnoea-hypopnea (AHI) scores indicative of sleep apnoea: 40% mild (AHI: 5 – 14), 26% moderate (AHI: 15 - 30) and 23% severe (AHI: > 30).

For our feasibility study in PLWD and their study partners, to date we have enrolled seven control participants (65 – 85 years) (3 females: 4 males) who had a mean age of 67.0 ± 6.2 years (range 61 – 79 years) and a mean BMI of 26.4 ± 3.8 kg/m^2^ (range: 22 – 32.6). We have enrolled 11 PLWD of whom eight (4 females: 4 males) have completed the study and six of these had study partners (3 females: 3 males). The PLWD had a mean age of 74.8 ± 4.4 years (range 67 – 81 years) and a mean BMI of 29.7 ± 7.6 kg/m^2^ (range: 20.7 – 42.1). The study partners had a mean age of 66.7 ± 14.8 years (range 44 – 80 years) and a mean BMI of 28.9 ± 4.1 kg/m^2^ (range: 23.9 – 33.6). These participants also report stable co-morbid medical conditions including type 2 diabetes, hypertension, arthritis and hypothyroidism, and are taking concomitant medications.

### Data completeness

The 35 cognitively intact participants from our initial study collected data for 7-14 days at home which combined was a total of 397 days/nights. We recorded from 6-10 devices (number of nights per device depended upon participant and device) which was a total of 2748 device days/nights from all the devices combined and we had 95% data completeness. In the laboratory, we recorded 35 nights with 8-10 devices which was 331 device nights, including PSG, and achieved 98% data completeness.

### Device acceptability

For all the wearable devices (n = 35 reported), participants rated the comfort as 3.8 ± 1.9 and the ease of use as 6.0 ± 1.2. For the nearable devices (n = 17 reported), participants rated the comfort as 6.3 ± 1.2 and the ease of use as 6.3 ± 1.1.

### Examples of at-home and in-laboratory recordings

One of the strengths of our approach and using multiple simultaneous devices is that it is possible to obtain concurrent physiological, behavioural, and environmental signals both at-home and in a laboratory setting. Here we show examples from individual participants of the datasets that can be obtained.

Figure 5 shows exemplar data from a cognitively intact male participant in their 60s who had moderate sleep apnoea (AHI = 24.1) and was living with controlled type 2 diabetes. The raster plot includes 14-consecutive days of home recording followed by the overnight in-laboratory session, simultaneously using two contactless, nearable technologies and a wrist worn actigraphy device (actigraphy was not used in the laboratory) with completion of a subjective sleep diary. The raster plot provides a measure of sleep behaviour and shows the day-to-day variation in sleep timing. The two under-mattress, contactless devices accurately detect bed presence as demonstrated by their concordance with the information captured in the sleep diary [52]. These devices can also capture daytime naps taken in bed and generate automated sleep summaries (boxed regions in the raster).

**Figure 5.**
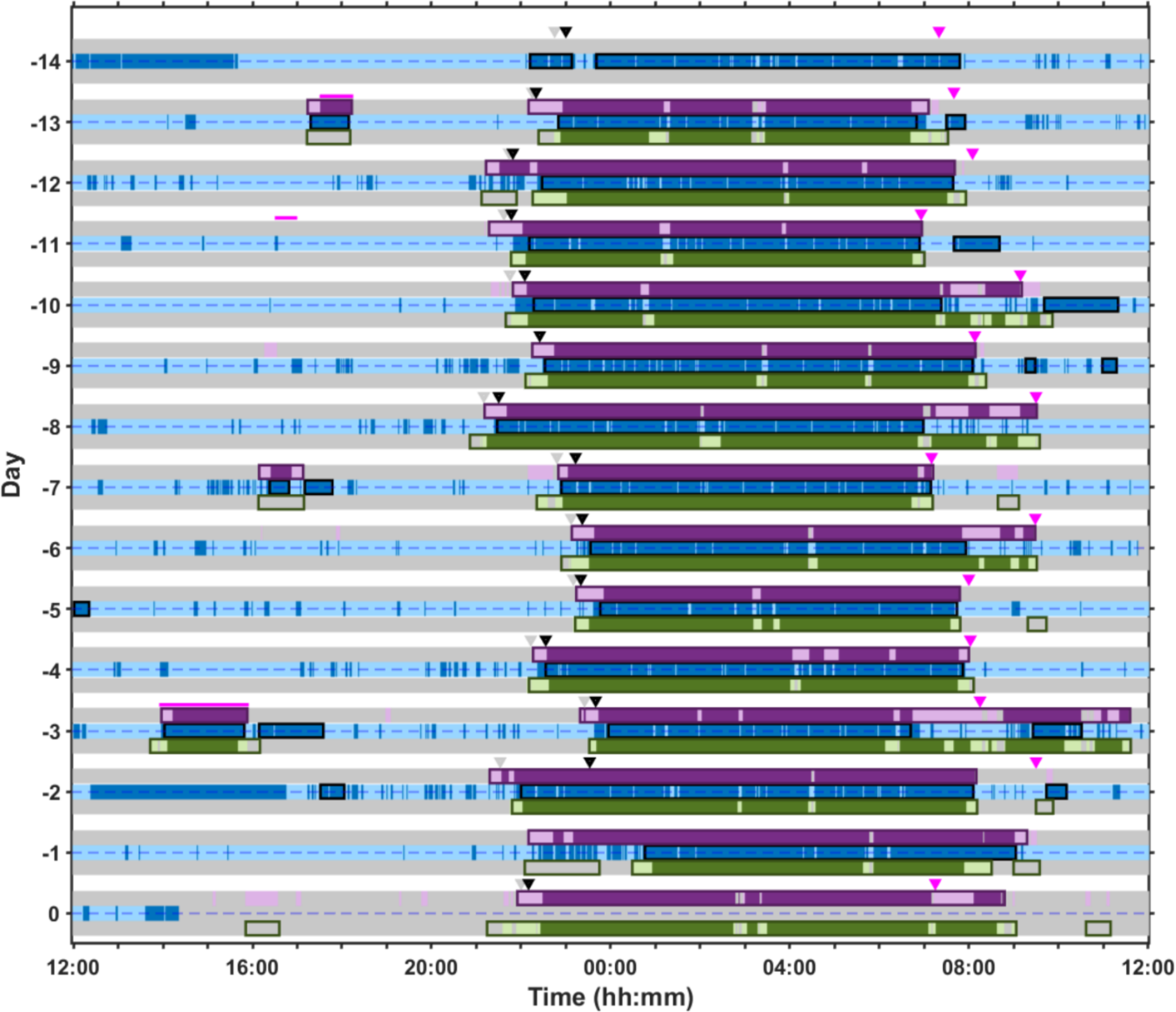
Multiple days of at-home recording (days -14 to -1) and a single overnight laboratory session (day 0) in a male participant in their 60s. The grey bars represent when the participant is out of bed and the purple and green bars represent when the participant is in bed as detected by two different ‘under the mattress’ nearable devices (Nearable 1 and nearable 2, respectively). For Nearable 1, the light pink represents periods of wake, and the darker purple represents sleep; for Nearable 2, the light green represents periods of wake and the darker green represents sleep. The bed entry and bed exit times recorded on the sleep diary are represented by inverted grey and pink triangles, respectively, and the black triangle indicates estimated sleep onset according to the sleep diary. The horizontal magenta lines represent nap times recorded on the sleep diary. The blue bars represent wrist worn actigraphy with dark blue indicating sleep and light blue representing wake.

Figure 6 provides an example of data captured in the laboratory session where polysomnography was recorded for 10 hours with simultaneous use of three nearable devices. For each nearable, discrepancy between the device and manually scored PSG are indicated with darker coloured bands at three different levels of sleep stage classification: 1) wake vs sleep, 2) wake vs NREM vs REM, 3) wake vs deep sleep vs light sleep vs REM. For all three nearable devices it can be seen that as the resolution of the sleep staging increases so do the number of discordant epochs. Thus, while the nearable devices might be able to distinguish between wake and sleep, they cannot accurately distinguish between different stages of sleep.

**Figure 6.**
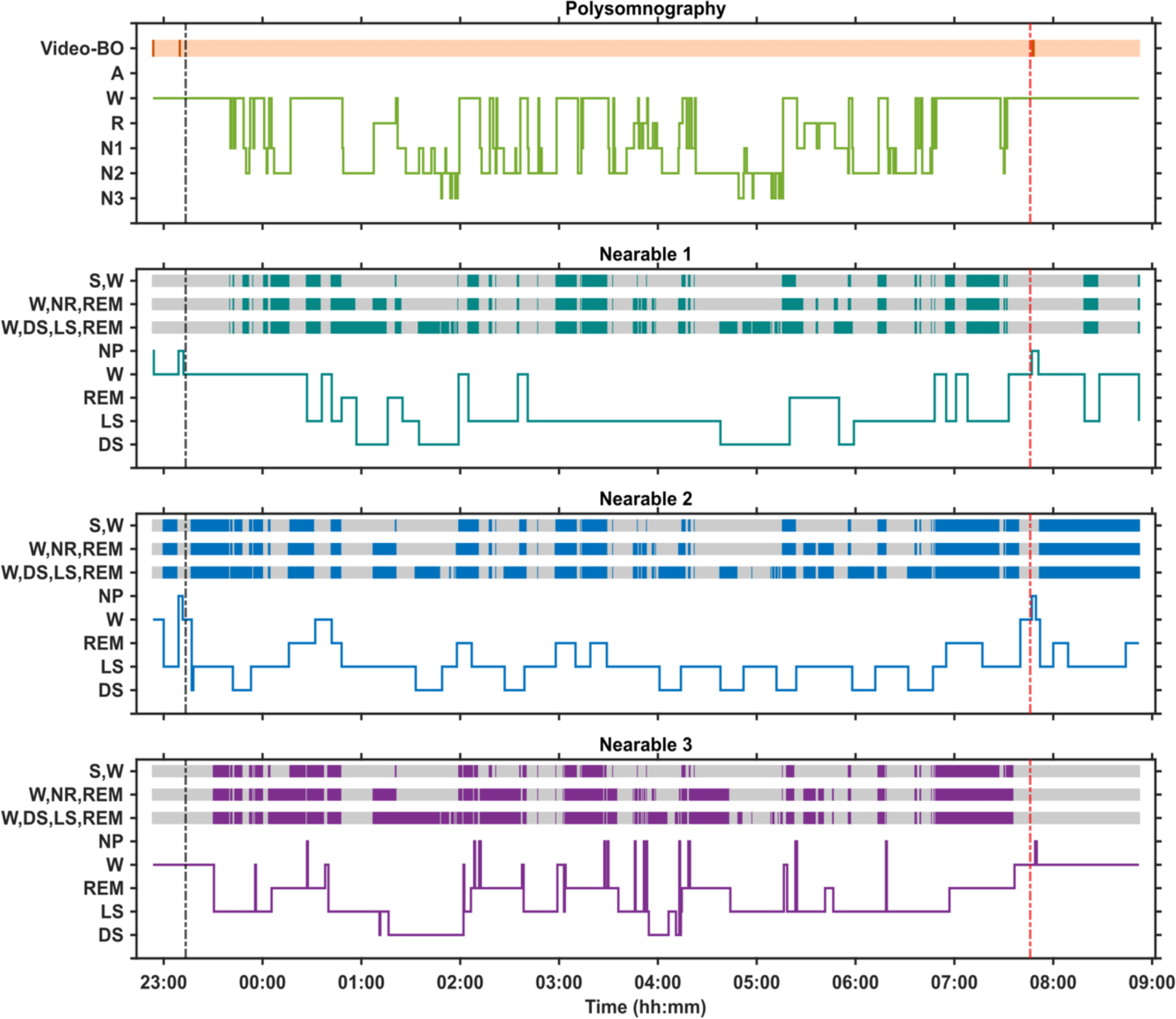
Hypnograms from a 10-hour in-bed period in a laboratory environment in a single participant with simultaneous polysomnography, including video, and three nearable devices. The black vertical dotted line depicts Lights Off and the red vertical dotted line depicts Lights On. The orange horizontal bar represents bed occupancy according to the video, with darker lines indicating when the participant left the bed. For each nearable, the discrepancy between the nearable and the PSG determined sleep is depicted at three different levels of sleep stage classification: 1) wake vs sleep, 2) wake vs NREM vs REM, 3) wake vs deep sleep vs light sleep vs REM. The darker coloured region indicates epochs of discrepancy. BO = bed occupancy, S = Sleep, W = wake, NR = NREM, NP = not present, REM = rapid eye movement sleep, LS = light sleep, DS = deep sleep.

Figure 7 provides an example of heart rate and breathing rate captured from PSG and three nearable devices from a single participant during a 10-hour period in bed in the laboratory. The pattern observed in the vital signs with Nearable 1 matches that detected by the gold standard PSG signals, whereas Nearable 2 records some spikes in heart rate that are not seen in the PSG or Nearable 1. Nearable 3 only records breathing rate, and the signal is consistent across the course of the night.

**Figure 7.**
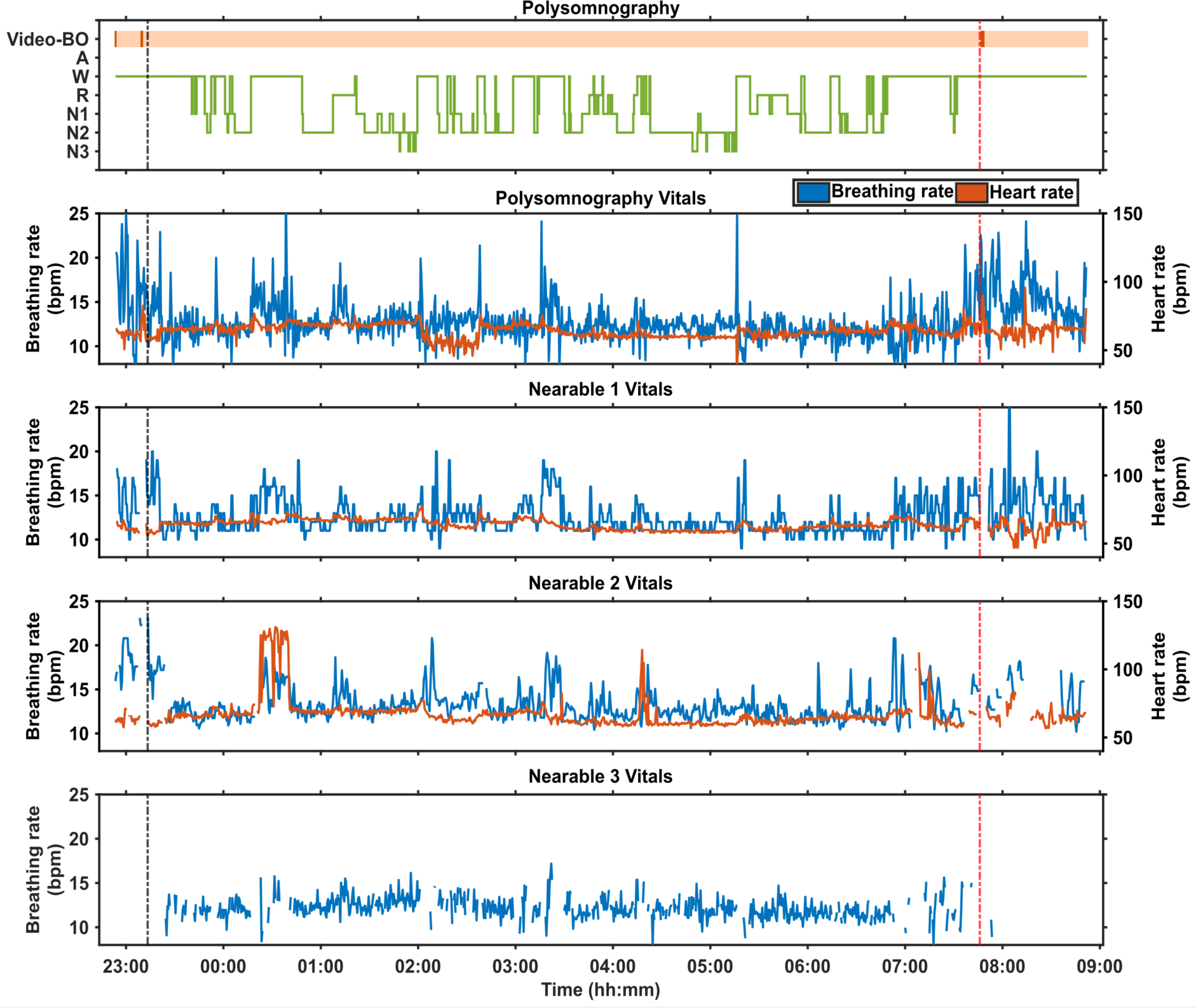
Physiological measures during a 10-hour in-bed period in a laboratory environment in a single participant with simultaneous recordings of polysomnography, including video, and from three nearable devices. The black vertical dotted line depicts Lights Off and the red vertical dotted line depicts Lights On. The orange horizontal bar represents bed occupancy according to the video, with darker lines indicating when the participant left the bed. For each nearable device, blue lines represent breathing rate and red lines represent heart rate. Within the PSG hypnogram, BO = bed occupancy, A = artefact, W = wake, R = REM sleep, N1 = stage 1 NREM sleep, N2 = stage 2 NREM sleep, N3 = stage 3 NREM sleep.

Figure 8 shows multiple consecutive days of recording in a woman living with dementia in her 70s and her partner, a man in his 80s, who share a bed. The PLWD’s sleep is fragmented and varies from day to day in terms of duration and timing. The partner also experiences nights of discontinuous sleep.

**Figure 8.**
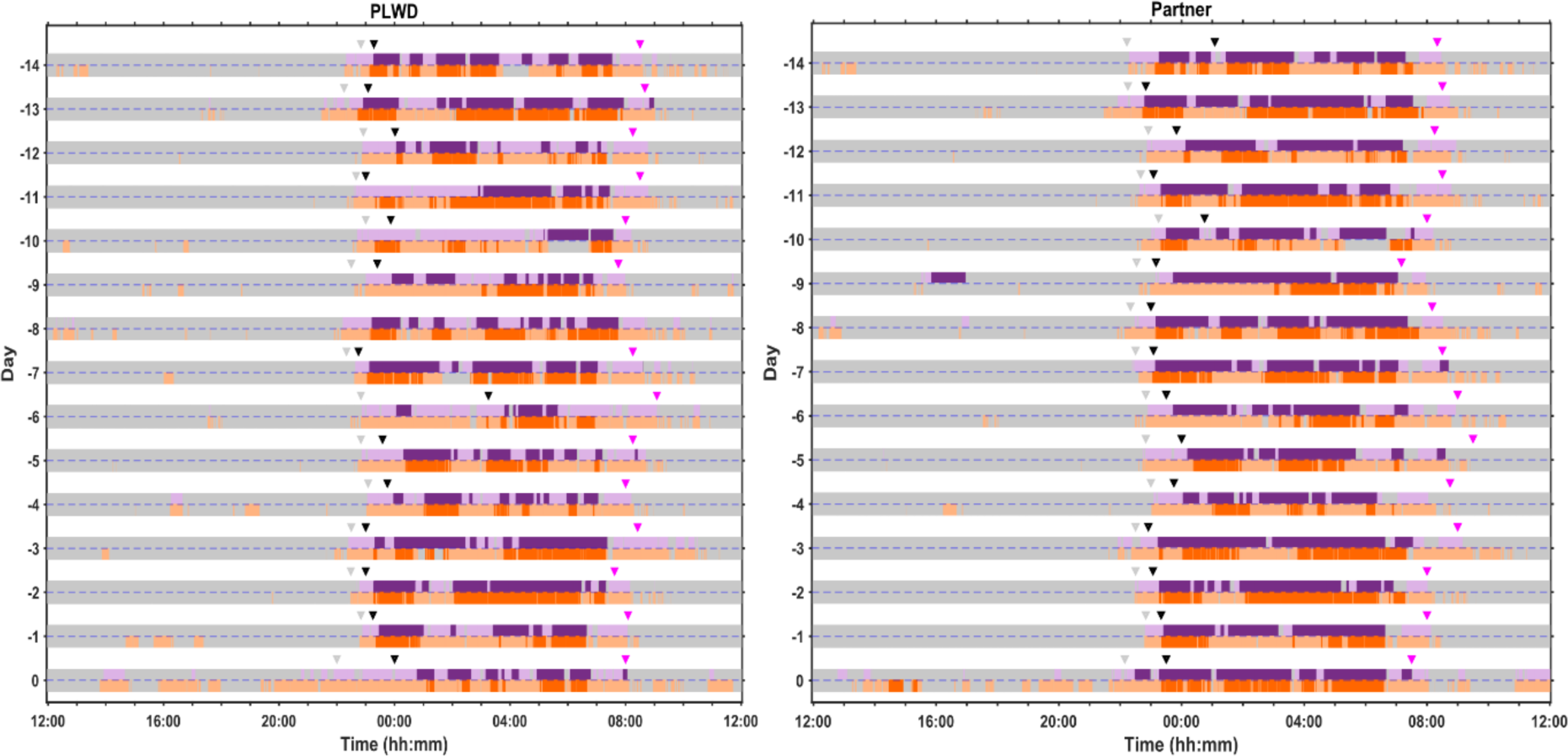
Multiple days of at-home recording (days -14 to -1) and a single overnight laboratory session (day 0) from a PLWD and their partner who share a bed. The grey bars represent when the participant is out of bed and the purple and red bars represent when the participant is in bed as detected by two different nearable devices (Nearable 1 and Nearable 2, respectively). The purple bars represent an under-mattress sensor and the red bars a bedside sensor; for both, the darker shading indicates sleep and the light shading indicates wake. The bed entry and bed exit times recorded on the sleep diary are represented by inverted grey and pink triangles, respectively, and the black triangle indicates estimated sleep onset according to the sleep diary.

Figure 9 is an example of light exposure data for a 24-hour period for an individual from two worn devices, as well as the light levels in the room in their house in which they spend the majority of their time. For the 17 participants who wore both the actiwatch and HOBO, the was a significant correlation between the measured values by the two devices (r = 0.36, p < 0.001). For this individual, their morning light exposure is up to 10,000 lux and exceeds the light levels in their home, suggesting they are outside. Their afternoon light exposure varies between 10 and 1000 lux but is generally lower than the light levels measured in their house suggesting they are indoors. Their evening light exposure is quite low and generally ≤ 100 lux.

**Figure 9.**
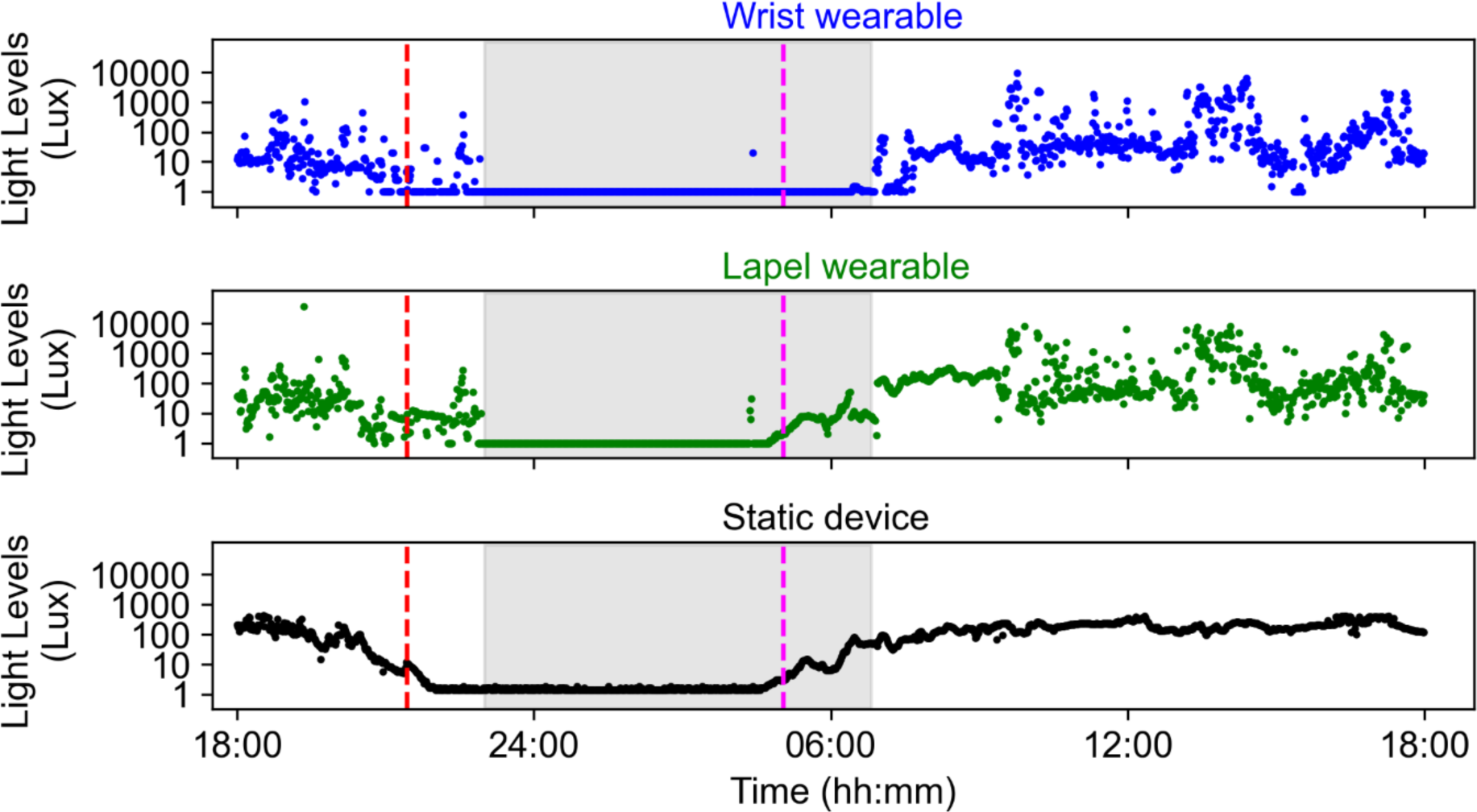
Light exposure data (plotted on a Log scale) measured over a 24-hour period for one participant using three devices: a wrist-worn wearable (blue), a collarbone/lapel worn wearable (green), and a static room device (black) (where the participant spends the majority of their time). Grey shaded areas indicate sleep periods reported in the sleep diary, the vertical red dashed line is sunset, and the vertical pink dashed line is sunrise.

## Discussion

We have described our successful approach for evaluating multiple, concurrent sleep/circadian monitoring technologies both at home and in the lab against accepted standard measures in older people and PLWD. This approach is rather different from published approaches in terms of the population enrolled, the number of devices evaluated simultaneously in one individual, the use of home and lab assessments, and the analysis interval used. Our high level of data completeness (95-98%) and participant retention (97%) is an indication of the success of our approach.

The majority of previous evaluation studies have taken the approach of either validating a single device against gold-standard or assessing multiple devices but not simultaneously in the same individual. For example, Chinoy and colleagues took a similar approach to us in simultaneously comparing seven consumer sleep tracking devices (wearable and contactless) in young participants at home and in the laboratory but participants only used a subset of the devices [48]. A strength of our design is that participants utilised multiple devices simultaneous to maximise evaluations and comparisons.

The predominant inclusion of young and healthy individuals in device evaluation studies limits the applications of the findings [48, 75–77]. In addition, only evaluating the performance of a device using self-selected lights off/on period when sleep efficiency is high means it is not possible to evaluate the ability of the device to discriminate quiet wake (e.g., [75, 76]. Indeed, in a young healthy population, it was demonstrated that the performance of both wearable and contactless devices worsened on a night of sleep disruption compared to an undisturbed night in the laboratory [48].

The value of assessing the device in the population in which it will be used was highlighted by three recent studies. An assessment of the EMFIT-QS mattress sensor in participants from a sleep disorders centre with a BMI of 33.8 ± 8.3 kg/m2 (mean ± SD; range 21.4 - 46.6) revealed that this device overestimated TST and underestimated WASO [78].

However, the authors noted that the performance actually worsened in those with a high apnoea-hypopnea index (AHI) and more fragmented sleep, but that estimations of TST actually improved in participants with increased weight and BMI suggesting that the device performs better with bigger movements from heavier individuals [78]. The Withings Sleep Mattress Analyser was assessed in participants with suspected sleep apnoea, and the device similarly overestimated sleep and underestimated wake but could accurately detect moderate to severe sleep apnoea (AHI was 31.2 ± 25 and 32.8 ± 29.9 with PSG and WSA, respectively) highlighting its diagnostic and monitoring value [79]. Finally, 293 cognitively normal and mildly impaired older adults were monitored for up to six nights at home using single channel EEG (scEEG), actigraphy and sleep diaries [80]. Estimates of TST showed the greatest agreement across the methods, particularly for cognitively intact adults, but the agreement between actigraphy and scEEG decreased in those with mild cognitive impairment and biomarker evidence of Alzheimer’s Disease. Thus, it is crucial that the device is evaluated in a relevant population for its intended use.

The devices we are testing do have potential for long-term use in the home environment. The Withings Sleep Analyser has been deployed in PLWD where variation in in night-time behaviour and physiology was shown to relate to disease progression, comorbid illnesses and changes in medication [81].

The approach we describe here could be applied to evaluate the performance and acceptability of any novel sleep or circadian monitoring device in any population. To best assess the device, it should be considered how the device would be used in the real-world and what information is most relevant e.g., bed occupancy, sleep structure, distribution of activity, light exposure. Our experimental design highlights the value of taking a multi- modal approach and assessing the performance of devices longitudinally at-home.

Combining information from multiple devices can assist with interpreting sleep-wake behaviour as well as allowing their performance to be cross-validated. For example, the AWS provides information about activity levels but not about whether the participant is in bed; however, when the AWS data is viewed in conjunction with the Withings Mattress data it is possible to identify when the participant has left the bed rather than just being restless in the bed. This is particularly relevant to PLWD whose nocturnal wandering is a major reason for them to be moved from their home into a care home.

One challenge of the protocol was that many participants had not used smart technology previously and we required participants to complete a number of procedures independently which some PLWD were concerned about whether they would remember. Thorough training sessions, provision of comprehensive written instructions, the role of the study partner for PLWD, and frequent contact between researchers and participants ensured that they felt supported and able to carry out all study procedures.

In conclusion, we have developed a novel protocol that allows multiple sleep/circadian/environmental technologies to be assessed simultaneously in an individual both at-home and in the laboratory. Our approach was successful in terms of data quality, data completeness, and gaining an understanding of device acceptability. The protocol was conducted in both cognitively intact older adults and PLWD to provide a comprehensive picture of an individual’s behaviour, physiology, and environment.

## Data Availability

The data used in this study are available from the corresponding author upon reasonable request.

## Acknowledgements

We thank Surrey Clinical Research Facility and Surrey and Borders Partnership Trust for support with recruitment and residential sessions and Daniel Barrett, Marta Messina, and Tegan Ward for their contribution to this work.

